# Omicron-associated changes in SARS-CoV-2 symptoms in the United Kingdom

**DOI:** 10.1101/2022.01.18.22269082

**Authors:** Karina-Doris Vihta, Koen B. Pouwels, Tim EA Peto, Emma Pritchard, Thomas House, Ruth Studley, Emma Rourke, Duncan Cook, Ian Diamond, Derrick Crook, David A. Clifton, Philippa C. Matthews, Nicole Stoesser, David W. Eyre, Ann Sarah Walker, the COVID-19 Infection Survey team

## Abstract

**Background:** The SARS-CoV-2 Delta variant has been replaced by the highly transmissible Omicron BA.1 variant, and subsequently by Omicron BA.2. It is important to understand how these changes in dominant variants affect reported symptoms, while also accounting for symptoms arising from other co-circulating respiratory viruses.

**Methods:** In a nationally representative UK community study, the COVID-19 Infection Survey, we investigated symptoms in PCR-positive infection episodes vs. PCR-negative study visits over calendar time, by age and vaccination status, comparing periods when the Delta, Omicron BA.1 and BA.2 variants were dominant.

**Results:** Between October-2020 and April-2022, 120,995 SARS-CoV-2 PCR-positive episodes occurred in 115,886 participants, with 70,683 (58%) reporting symptoms. The comparator comprised 4,766,366 PCR-negative study visits (483,894 participants); 203,422 (4%) reporting symptoms. Symptom reporting in PCR-positives varied over time, with a marked reduction in loss of taste/smell as Omicron BA.1 dominated, maintained with BA.2 (44%/45% 17 October 2021, 16%/13% 2 January 2022, 15%/12% 27 March 2022). Cough, fever, shortness of breath, myalgia, fatigue/weakness and headache also decreased after Omicron BA.1 dominated, but sore throat increased, the latter to a greater degree than concurrent increases in PCR-negatives. Fatigue/weakness increased again after BA.2 dominated, although to a similar degree to concurrent increases in PCR-negatives. Symptoms were consistently more common in adults aged 18-65 years than in children or older adults.

**Conclusions:** Increases in sore throat (also common in the general community), and a marked reduction in loss of taste/smell, make Omicron harder to detect with symptom-based testing algorithms, with implications for institutional and national testing policies.

**Summary:** In a UK community study, loss of taste/smell was markedly less commonly reported with Omicron BA.1/BA.2 than Delta SARS-CoV-2 infections, with smaller declines in reported shortness of breath, myalgia and fatigue/weakness, but increases in sore throat, challenging symptom-based testing algorithms.

## Introduction

As the highly-transmissible SARS-CoV-2 Omicron variants BA.1 and BA.2 have emerged and become dominant, coincident with other winter respiratory viruses circulating in the Northern hemisphere, changes in symptomatology may influence clinical and testing policy. Experimental and clinical data suggest Omicron has less impact on the lower respiratory tract, leading to less severe disease[1–7], with the variant-defining mutations potentially also affecting other symptoms.

We used the UK Covid-19 Infection Survey, a nationally representative longitudinal household study[8], to investigate if SARS-CoV-2 infection symptoms have changed with the Omicron variants. We compared the probability of reporting any symptoms, as well as the probability of reporting specific symptoms in both SARS-CoV-2 PCR-positive infection episodes and comparator PCR-negative study visits by calendar time, vaccination status and age. We focused on comparisons between time periods when the Delta variant (described previously only to August 2021[9]), Omicron BA.1 and Omicron BA.2 were dominant in the UK[10].

## Methods

This analysis was based on SARS-CoV-2 PCR tests of nose and throat swabs from 1 October 2020 to 23 April 2022 in the Office for National Statistics (ONS) Covid Infection Survey (CIS) (ISRCTN21086382, https://www.ndm.ox.ac.uk/covid-19/covid-19-infection-survey/protocol-and-information-sheets). The survey randomly selects private households on a continuous basis from address lists and previous surveys to provide a representative UK sample.

Following verbal agreement to participate, a study worker visited each household to take written informed consent, which was obtained from parents/carers for those 2-15 years; those aged 10-15 years provided written assent. Those <2 years were not eligible, to avoid asking parents to swab babies and very young children. Ethical approval was provided by the South Central Berkshire B Research Ethics Committee (20/SC/0195).

Individuals were asked about demographics, symptoms, contacts and relevant behaviours (https://www.ndm.ox.ac.uk/covid-19/covid-19-infection-survey/case-record-forms). To reduce transmission risks, participants ≥12 years self-collected nose and throat swabs following study worker instructions. Parents/carers took swabs from children 2-11 years. At the first visit, participants were asked for consent for optional follow-up visits every week for the next month, then monthly from enrolment. While participants were offered the option of a single visit, 99% of participants participated in longitudinal sampling (**Table S1**).

Swabs were analysed at the UK’s national Lighthouse Laboratories at Milton Keynes and Glasgow using identical methodology. PCR for three SARS-CoV-2 genes (N protein, S protein and ORF1ab) was performed using the Thermo Fisher TaqPath RT-PCR COVID-19 kit, and analysed using UgenTec FastFinder 3.300.5, with an assay-specific algorithm and decision mechanism that allows conversion of amplification assay raw data from the ABI 7500 Fast into test results with minimal manual intervention. Samples are called positive if at least the N gene and/or ORF1ab are detected. Although S gene cycle threshold (Ct) values are determined, S gene detection alone is not considered sufficient to call a sample positive.

The presence of 12 specific symptoms in the previous seven days was elicited at each visit from the start of the survey (cough, fever, myalgia, fatigue/weakness, sore throat, shortness of breath, headache, nausea, abdominal pain, diarrhoea, loss of taste, loss of smell), as was whether participants thought they had (unspecified) symptoms compatible with COVID-19. Positive response to any of these questions defined “symptomatic” cases. Four additional symptoms (runny nose, trouble sleeping, loss of appetite, wheezing) were added from 29 September 2021; as these were not elicited throughout the survey, they were considered separately and not used to define symptomatic cases.

We grouped repeated PCR-positive tests into infection “episodes”[11], and included the first positive study test in each episode in analysis (details in Supplementary Methods). Each positive episode was characterised as wild-type/Delta/Omicron BA.2-compatible if the S-gene was ever detected (by definition, with N/ORF1ab/both), or as Alpha- or Omicron BA.1-compatible if positive at least once for ORF1ab+N (and never for the S-gene), otherwise “other” (N-only/ORF1ab-only) depending on calendar period (**Fig.1A**). Symptom presence was defined as reported symptoms at any visit within [0,+35] days of the first PCR-positive test in each infection episode (i.e. spanning [-7,+35] days given the question timeframe), to allow for the random sampling leading to pre-symptomatic identification of some individuals, who only reported symptoms subsequently.

**Figure 1.**
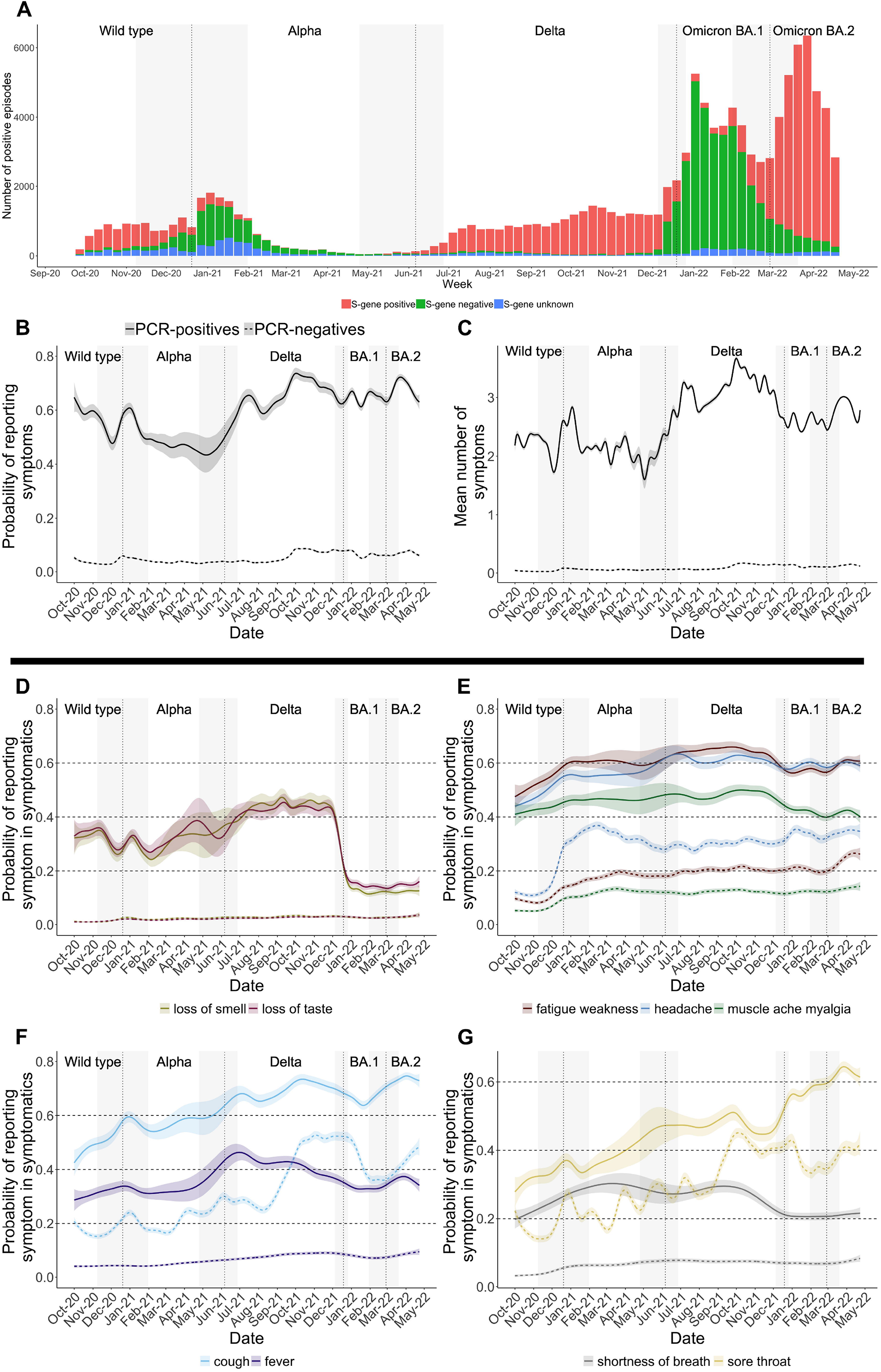
(A) Variants and (B)-(G) symptoms in those testing positive and negative for SARS-CoV-2 over time in the UK. Panel A shows the number of PCR-positive infection episodes that were S-gene negative (Alpha-compatible 20 December 2020 to 5 June 2021; Omicron BA.1-compatible 19 December 2021 to 26 February 2022) and S-gene positive (Delta-compatible 6 June 2021 to 18 December 2021; Omicron BA.2-compatible from 27 February 2022 onwards). Vertical lines indicate periods when new variants came to dominate based on gene positivity patterns (>50% of PCR-positives): wild type before 20 December 2020, then Alpha before 5 June 2021, then Delta before 19 December 2021 then Omicron BA.1 before 27 February 2022; Omicron BA.2 became the dominant variant afterwards. Panels B and C show the probability of reporting symptoms and the number of symptoms (out of the 12 elicited throughout the study period) of all PCR-positive infection episodes and all PCR-negative comparator visits. Panels D-G show the probability of specific symptoms in symptomatic PCR-positive infection episodes and in symptomatic PCR-negative comparator study visits, after adjustment for age, sex, ethnicity (presented at the reference category age 45, male, white).

As a comparator, we initially considered all visits with negative PCR tests, and then, following a previous analysis to August 2021[9], excluded visits where symptoms could plausibly be related to ongoing effects of COVID-19 or long COVID, where there was a high pre-test probability of a new COVID-19 infection that had not been detected in the study, or where symptoms were likely driven by recent vaccination (details in Supplementary Methods).

Generalised additive models (binomial distribution with complementary log-log link, R mgcv (v.1.8-31) package) were fitted to estimate the percentage of PCR-positive infection episodes and PCR-negative visits that were symptomatic, and the percentage of symptomatic PCR-positive infection episodes and symptomatic PCR-negative visits reporting each symptom separately. Models adjusted simultaneously for calendar time (smoothing spline), age (smoothing spline), sex and ethnicity (white vs non-white). To explore differences between Delta, Omicron BA.1, and Omicron BA.2 infections by vaccination status and infection/re-infection, we restricted PCR-positives to those occurring after 29 September 2021 and classified S-gene negatives occurring after 1 December 2021 as Omicron BA.1-compatible (34,576 infections, 20,345 [59%] symptomatics), and S-gene positives 29 September 2021-2 January 2022 as Delta-compatible (14,318 infections, 9,030 [63%] symptomatics) and 30 January 2022-23 April 2022 as Omicron BA.2-compatible (34,796 infections, 22,591 [65%] symptomatics) (excluding S-gene positives 3-29 January 2022 as both Delta and Omicron BA.2 infections occurred during this period and genetic sequences were not available for all PCR-positives).

## Results

Between October 2020 and April 2022, 120,995 PCR-positive episodes occurred in 115,886 participants (median 44 years, IQR 24-61), 70,683 (58%) with reported symptoms. 8898/120,995 (7%) were re-infections (**Fig.S1**), 4244 (48%) with reported symptoms. The comparator comprised 4,766,366 PCR-negative study visits (483,894 participants, median 55 years, IQR 36-68); 203,422 (4%) with reported symptoms.

While Omicron BA.1 infections dominated (19 December 2021 to 26 February 2022, when >50% of PCR-positive results were S-gene negative), the percentage of PCR-positive infection episodes with reported symptoms was lower compared to much of the previous time period when the Delta variant dominated (6 June 2021 to 18 December 2021, **Fig.1B/C**). Reporting any symptoms increased again after Omicron BA.2 became the dominant variant (27 February 2022 onwards, when >50% of PCR-positive results were S-gene positive). For both Omicron BA.1 and BA.2 the mean number of symptoms reported in PCR-positive infection episodes was lower than with Delta, but was higher with BA.2 than BA.1. Changes in the percentage reporting any symptoms at PCR-negative visits, and the mean number of symptoms reported at PCR-negative visits, were much smaller over these time periods, with very slight increases from October 2021 onwards likely due in part to other seasonal infections.

For specific symptoms, amongst symptomatic PCR-positive infection episodes, there was a marked decline in reported loss of taste/smell for both Omicron variants, BA.1 and BA.2, from high levels during the period when Delta dominated, e.g. from 44%/45% on 17 October 2021 (approximately peak Delta, **Fig.1A**), to 16%/13% on 2 January 2022 (approximately peak BA.1) with only very small changes thereafter, e.g. to 15%/12% on 27 March 2022 (approximately peak BA.2). Although loss of taste/smell was also more uncommon with Alpha than Delta, it was even more uncommon with Omicron BA.1/BA.2 than Alpha (**Fig.1D**). Loss of taste/smell remained extremely uncommon in symptomatic PCR-negative visits throughout (**Fig.1D**).

There were concurrent smaller, but significant, declines in symptomatic PCR-positive infection episodes with reported cough, fever, fatigue/weakness, myalgia, shortness of breath and headache during December 2021, as Omicron BA.1 dominated (**Fig.1E/F/G**). As Omicron BA.2 became dominant, cough and to a lesser extent fever and fatigue/weakness increased again, while shortness of breath, myalgia, and headache remained at similar levels to those observed with BA.1 (**Fig.1E/F/G**). The main changes in the percentages of symptomatic PCR-negative visits where these specific symptoms were reported was a substantial increase in cough in October 2021, which then decreased in January 2022 from 52% to 36%, before increasing again to 48% by 23 April 2022 (**Fig.1G**), and increases in headache over December 2021 (from 30% to 35%) and in fatigue/weakness over March 2022 (from 20% to 26%) (**Fig.1E**).

In contrast to these declines in other symptoms as Omicron BA.1 dominated, sore throat became more commonly reported with BA.1 and increased further with BA.2, from 46% to 56% in symptomatic PCR-positive infection episodes during December 2021, increasing further to 64% by April 2022. Similarly to cough, sore throat became more commonly reported at PCR-negative visits during October 2021, if anything dropping slightly in January 2022 from 43% to 33% before increasing again to 42% by 23 April 2022 (**Fig.1G**). These changes were smaller in symptomatic PCR-negatives than symptomatic PCR-positives, i.e., were insufficient to explain Omicron-associated increases in sore throat.

Gastrointestinal symptoms were reported infrequently in symptomatic PCR-positive infection episodes regardless of variant, and were reported at similar frequencies at PCR-negative visits (**Fig.S2**). Reporting of runny nose generally followed reporting of sore throat, whereas other symptoms generally declined with Omicron BA.1/BA.2 (**Fig.S2**).

Differential symptom reporting between variants from 29 September 2021, particularly fewer cases with loss of taste/smell and more with sore throat, was broadly unaffected by vaccination status (13,317 (16%), 2,919 (4%), 16,459 (20%) and 50,314 (61%) of PCR-positive infection episodes occurred in those unvaccinated or vaccinated once, twice or three times respectively; full split by variant and evidence of symptoms in **Table S2**) (**Fig.2**). Similarly, changes in symptoms by variant were also relatively unaffected by whether the PCR-positive infection episode was the first infection (91%) vs. reinfection (9%) (**Fig.3**). However, overall, symptoms were less commonly reported in subsequent infections occurring from 29 September 2021 onwards (50%), compared to first infections during this time period (63%), but specific symptoms were reported at broadly similar frequencies in participants who were symptomatic in PCR-positive first and subsequent infections with Delta and Omicron BA.1 and BA.2 variants.

**Figure 2.**
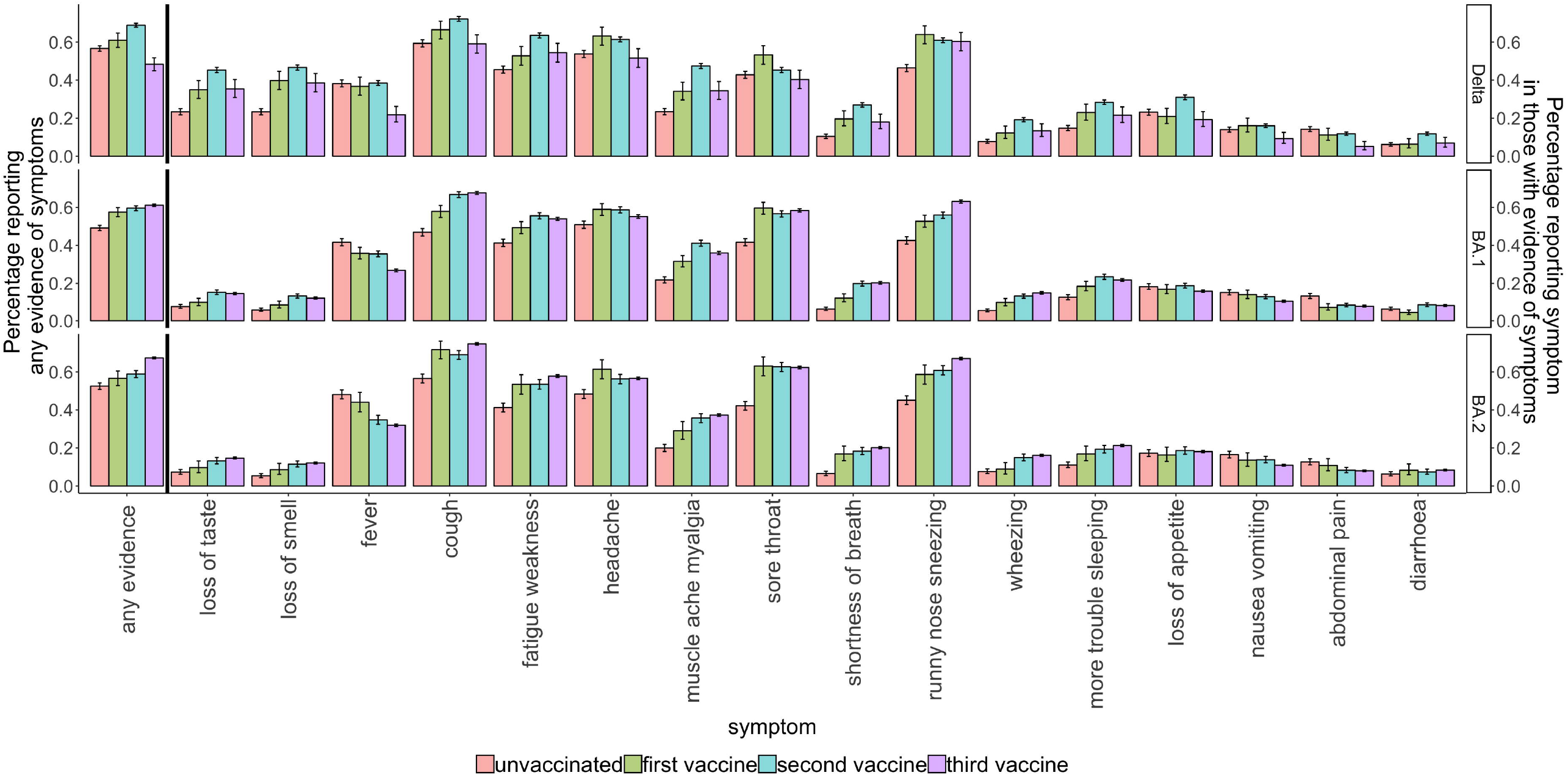
Percentage of PCR-positives reporting any evidence of symptoms, and percentage of symptomatic PCR-positives reporting specific symptoms from 29 September 2021 onwards by variant and by vaccination status. Note: not adjusted for other factors, see **Fig.4** for adjusted effect of age. Unvaccinated=before first vaccination at index positive test or never vaccinated, first vaccine= 21 days after first vaccination to 13 days after second, second vaccine=14 days after second vaccination to 13 days after third; third vaccine=14 days after third vaccination to 13 days after fourth. Fourth vaccination data not shown as less than 100 infections with evidence of symptoms (**Table S2**).

**Figure 3.**
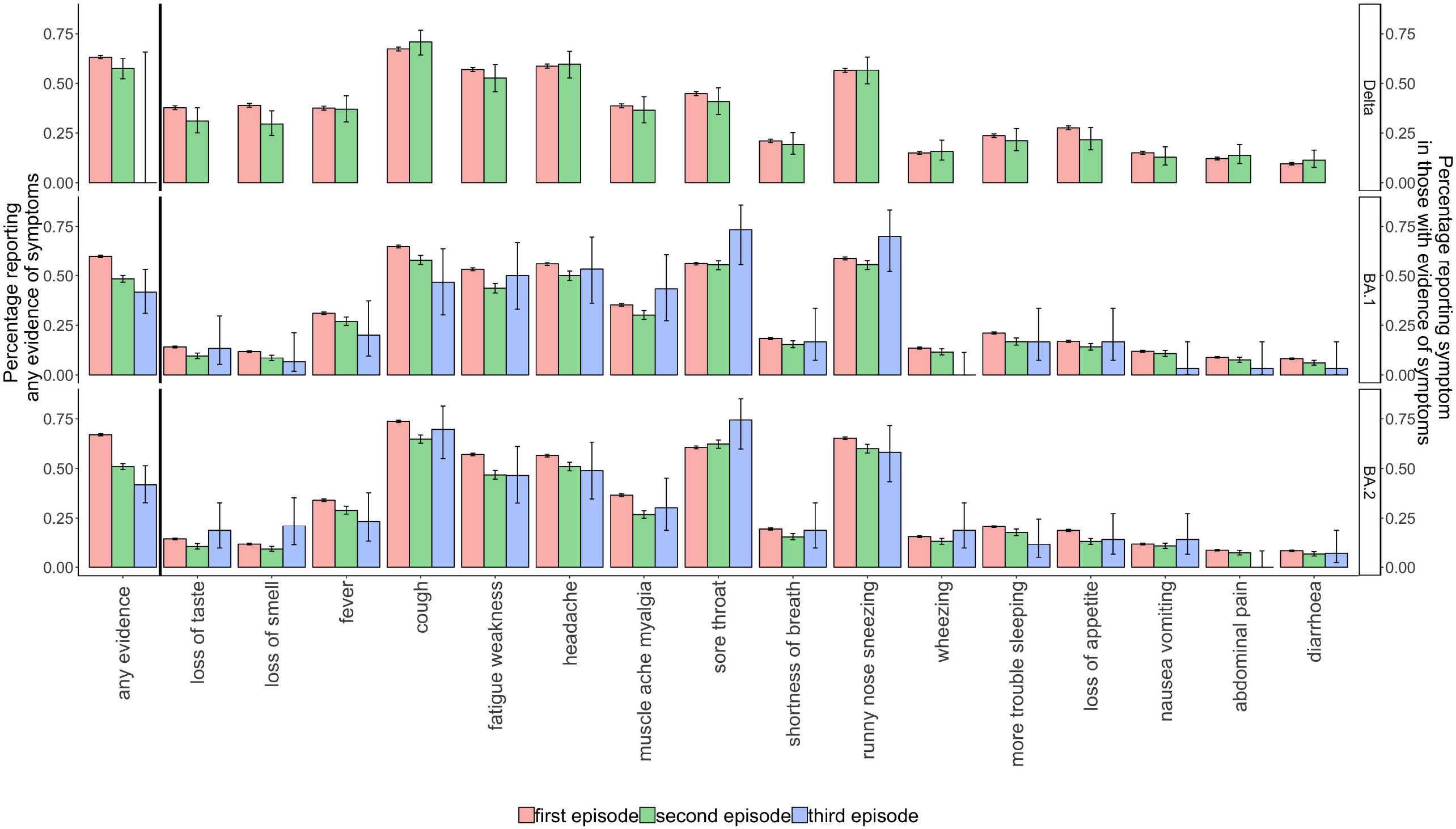
Percentage of PCR-positives reporting any evidence of symptoms, and percentage of symptomatic PCR-positives reporting specific symptoms from 29 September 2021 onwards by variant and infection/re-infection. Note: not adjusted for other factors, see **Fig.4** for adjusted effect of age.

There were differences in reported symptoms with these different variants by age when comparing reported symptoms at the peaks of the Delta, BA.1 and BA.2 waves (**Fig.4**). Adults aged 18-65 years were more likely to report the presence of any symptoms than children or adults >65 years. There was generally no evidence of difference in reporting the presence of any symptoms between Delta and BA.2, but there was a lower probability of reporting any symptoms with BA.1 across most ages. However, the mean number of symptoms reported with both BA.1 and BA.2 were generally lower across the ages compared to Delta, with the exception of the youngest and oldest for which there was no evidence of difference in the mean number of symptoms between BA.1 and Delta, but a higher mean number of symptoms for BA.2 vs Delta. Symptoms were less likely to be reported in PCR-positive infection episodes in children than younger adults, even more so with Omicron BA.1 than Delta infections and BA.2 (**Fig.S4**), whereas symptoms were most likely to be reported at PCR-negative visits in children, in particular cough and fever.

**Figure 4.**
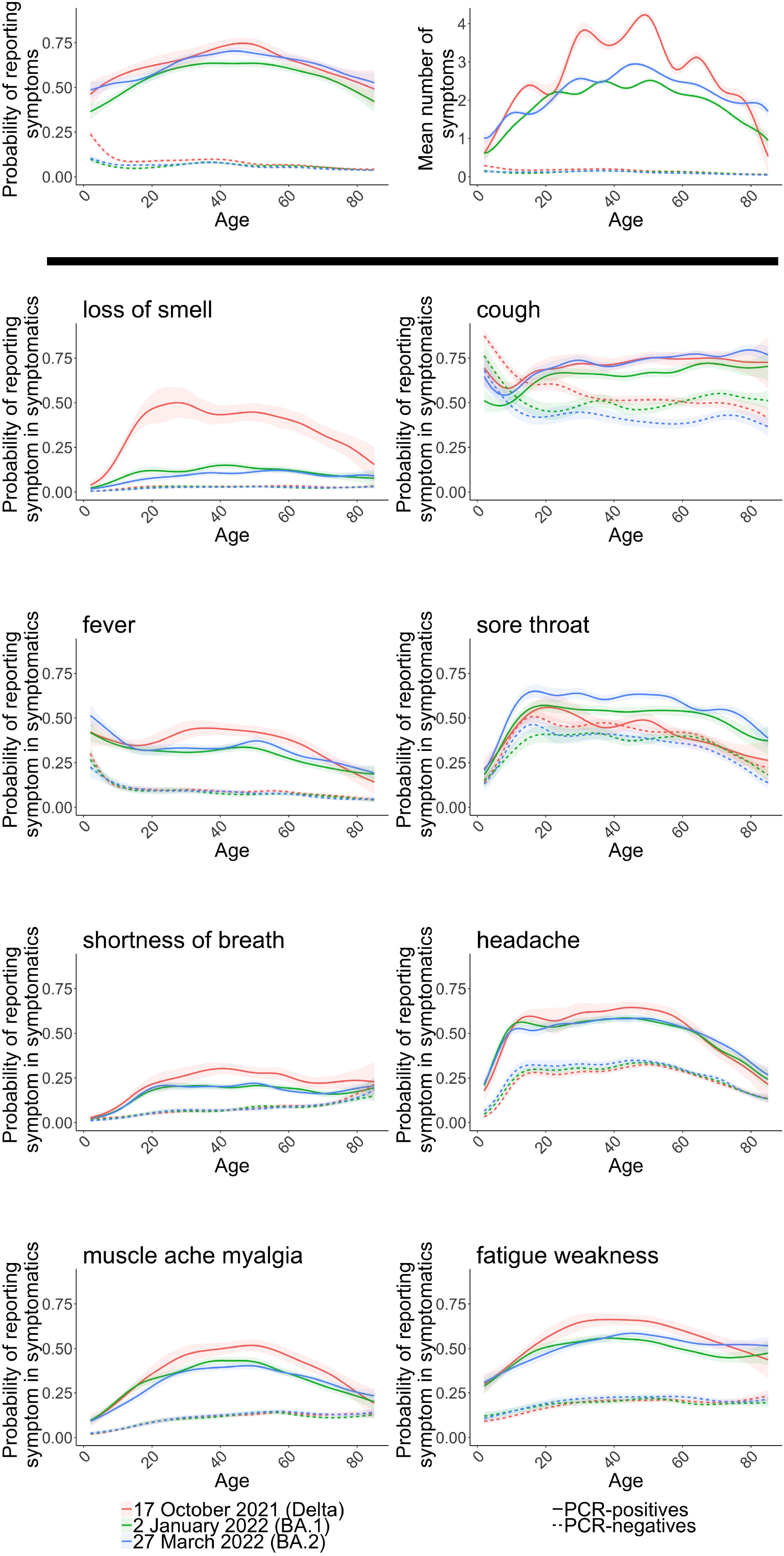
By age, estimated percentage of PCR-positives infection episodes and comparator PCR-negative study visits reporting symptoms and mean number of symptoms, and percentage of symptomatic PCR-positives and symptomatic PCR-negatives reporting specific symptoms, on 17 October 2021 (Delta), 2 January 2022 (when Omicron BA.1-compatible infections represented the highest proportion of PCR-positives), and 27 March 2022 (when Omicron BA.2 was the dominant variant). The panels in the first row show the probability of reporting symptoms and the number of symptoms (out of the 12 elicited throughout the study period) in all PCR-positive infection episodes and all PCR-negative comparator visits from 29 September 2021 onwards, estimated at three reference categories, 17 October 2021, 2 January 2022, and 27 March 2022. The remaining panels show the probability of reporting specific symptoms in symptomatic PCR-positive infection episodes and in symptomatic PCR-negative comparator study visits at these reference categories. All are adjusted for calendar date, age (allowing for effect modification by calendar date by including an interaction between calendar date and age), sex (reference category male), ethnicity (reference category white). See **Fig.S3** for other symptoms.

Loss of taste or smell was most commonly reported with Delta infections in adults aged 18-70 years, but at lower levels in older adults, and rarely in younger children; it was only seen at low levels regardless of age with Omicron BA.1/BA.2 infections. Variations in the percentage of symptomatic participants reporting most other specific symptoms across ages were broadly similar before vs after Omicron BA.1 dominated, but slightly higher percentages of symptomatic PCR-positive infection episodes in participants over 70 years reported fever, headache, fatigue/weakness and muscle ache/myalgia after Omicron BA.1/BA.2 dominated (**Fig.4**). Most specific symptoms were reported less frequently at infections in young children than adolescents/young adults regardless of the dominating variant, excepting fever which was reported significantly more with Omicron BA.1 and BA.2 infections in young children than adolescents/young adults, particularly for BA.2 (**Fig.S4**).

The net result of changes in the symptom profile, overall and by age, was that fever and cough became most strongly associated with PCR-positivity in those reporting symptoms after Omicron BA.2 became dominant, adjusting for age, sex and ethnicity (see Supplementary Methods) (**Fig.S5**). Although far less strongly associated than during the period when Delta was the main variant, loss of taste was still the fourth most strongly associated symptom after Omicron BA.2 dominated, with fatigue/weakness also strongly associated. These same four symptoms were also most strongly associated with PCR-positivity when Omicron BA.1 dominated. Sore throat was positively associated with PCR-positivity during the BA.2 dominant period, and to a slightly lesser degree with PCR-positivity during the BA.1 dominant period, while in contrast, sore throat was less likely to occur in symptomatic PCR-positives compared to symptomatic PCR-negatives in the Delta period.

## Discussion

In this study of predominantly mild community-based infection, overall Omicron BA.1 and BA.2 were associated with less loss of taste, loss of smell, shortness of breath, myalgia, fatigue/weakness and headache, but more sore throat, compared with Delta. The overall probability of reporting any symptoms was similar for Delta and BA.2, but lower for BA.1 regardless of age, while the mean number of symptoms reported was generally lower for both BA.1 and BA.2 compared to Delta across ages, although higher overall for BA.2 than BA.1. However, this was driven by symptomatology in adults; in the youngest and oldest participants, there was no evidence of difference in the percentage reporting any symptoms between BA.2 and Delta, and a higher mean number of symptoms were reported with BA.2 in the very youngest and oldest compared to both BA.1 and Delta.

In PCR/lateral flow antigen-positive cases, the ZOE study, which relies on volunteers reporting symptoms daily using an app, found a lower median number of symptoms reported in infections from 28 November 2021 to 17 January 2022 (predominantly Omicron BA.1) than 1 June to 27 November 2021 (predominantly Delta) matched by age, sex and ethnicity in those who had had a second or third vaccine[12], with less loss of smell and more sore throat being reported with Omicron BA.1, as in our study. The major strength of our study is that regular PCR testing was undertaken in all participants at all visits irrespective of symptoms. This provides a representative sample of PCR-negative visits without SARS-CoV-2 infection for comparison with symptom rates in PCR-positives. This is important because some symptoms reported in PCR-positive infections could be due to co-infections with other circulating respiratory viruses. Therefore, although our study does not specifically test for other viruses, we can estimate whether changes seen with Omicron BA.1 and BA.2 differ from underlying trends in the general population (**Fig.1D-G**), supporting much of the increase in sore throat being attributable to Omicron rather than other infections. We are also able to demonstrate large shifts in symptoms reported at PCR-negative visits over time, with concurrent increases in cough and sore throat in October 2021 likely reflecting other respiratory viruses. We also note that the probability of reporting any symptoms as well as specific symptoms, varied considerably during the periods when specific variants dominated, potentially reflecting how the survey captures more infections earlier on when positivity is rising, and more later on as positivity is decreasing[13]. We compared rates at the peak of each dominating variant to capture similar phases of the epidemic, as well as considering how these changed over time.

Intriguingly, we found that the differences between variants in the probability of reporting specific symptoms in symptomatic PCR-positives persisted regardless of vaccination status or whether the infection was the first or subsequent, while the probability of reporting symptoms was smaller for reinfections compared to first infections.

Limitations of our study include the fact that we cannot have certainty in determining reinfections given the data available; however, estimated reinfections were infrequent (7%), even once Omicron dominated (11%) and symptom profiles were broadly similar in first and subsequent infections from 29 September 2021. Another limitation is that the study does not collect data on healthcare provider visits, hospitalizations, or death, to allow analysis of the severity of Omicron infections beyond reported symptoms. The ZOE study found lower self-reported hospitalisation rates with infections occurring during the Omicron BA.1-dominant vs Delta-dominant period, and shorter duration of symptoms[12], and several other studies have documented lower hospitalisation rates with Omicron BA.1[14–17].

Increases in sore throat (also commonly reported at symptomatic PCR-negative visits), and the marked reduction in the previously highest specificity symptoms, namely loss of taste/smell, present challenges for testing algorithms. Previously during periods when wild-type, Alpha and Delta variants dominated, fever, cough or loss of taste/smell have been shown to offer a good balance between sensitivity and specificity for detecting SARS-CoV-2 infections[9]. In the UK, for much of the pandemic to date, any of these four symptoms formed a basis for the general public accessing PCR testing. However, changes in symptoms with Omicron mean that symptom-based screening for testing is now much more difficult, and have resulted in much broader criteria for symptoms suggestive of COVID being proposed[18], albeit with likely decreased specificity. In conclusion, changes in SARS-CoV-2 infection symptoms mean that Omicron is harder to detect with symptom-based testing algorithms with implications for institutional and national testing policies.

## Data Availability

Data are still being collected for the COVID-19 Infection Survey. De-identified study data are available for access by accredited researchers in the ONS Secure Research Service (SRS) for accredited research purposes under part 5, chapter 5 of the Digital Economy Act 2017. For further information about accreditation, contact or visit the SRS website.

## Declarations

### Contributors

This specific analysis was designed by ASW, K-DV, KBP, PCM, NS, DWE, TH, DC, TEAP. K-DV conducted the statistical analysis of the survey data. K-DV, NS, PCM, ASW drafted the manuscript. All authors contributed to interpretation of the study results, and revised and approved the manuscript for intellectual content. K-DV is the guarantor and accepts full responsibility for the work and conduct of the study, had access to the data, and controlled the decision to publish. The corresponding author (K-DV) attests that all listed authors meet authorship criteria and that no others meeting the criteria have been omitted.

### Ethical approval

The study received ethical approval from the South Central Berkshire B Research Ethics Committee (20/SC/0195).

### Transparency

The lead authors affirm that the manuscript is an honest, accurate, and transparent account of the study design being reported, no important aspects of the study have been omitted, and any discrepancies from the study as originally planned (and, if relevant, registered) have been explained. Dissemination to participants and related patient and public communities: Results of individual tests were communicated to the participants. Overall study results were disseminated through the preprint of the study. Findings were disseminated in lay language in the national and local press.

## Acknowledgements

We are grateful for the support of all COVID-19 Infection Survey participants and the COVID-19 Infection Survey team:

Office for National Statistics: Sir Ian Diamond, Emma Rourke, Ruth Studley, Tina Thomas, Duncan Cook.

Office for National Statistics COVID Infection Survey Analysis and Operations teams, in particular Daniel Ayoubkhani, Russell Black, Antonio Felton, Megan Crees, Joel Jones, Lina Lloyd, Esther Sutherland.

University of Oxford, Nuffield Department of Medicine: Ann Sarah Walker, Derrick Crook, Philippa C Matthews, Tim Peto, Emma Pritchard, Nicole Stoesser, Karina-Doris Vihta, Jia Wei, Alison Howarth, George Doherty, James Kavanagh, Kevin K Chau, Stephanie B Hatch, Daniel Ebner, Lucas Martins Ferreira, Thomas Christott, Brian D Marsden, Wanwisa Dejnirattisai, Juthathip Mongkolsapaya, Sarah Cameron, Phoebe Tamblin-Hopper, Magda Wolna, Rachael Brown, Sarah Hoosdally, Richard Cornall, Yvonne Jones, David I Stuart, Gavin Screaton.

University of Oxford, Nuffield Department of Population Health: Koen Pouwels.

University of Oxford, Big Data Institute: David W Eyre, Katrina Lythgoe, David Bonsall, Tanya Golubchik, Helen Fryer.

University of Oxford, Radcliffe Department of Medicine: John Bell.

Oxford University Hospitals NHS Foundation Trust: Stuart Cox, Kevin Paddon, Tim James. University of Manchester: Thomas House.

Public Health England: John Newton, Julie Robotham, Paul Birrell.

IQVIA: Helena Jordan, Tim Sheppard, Graham Athey, Dan Moody, Leigh Curry, Pamela Brereton.

National Biocentre: Ian Jarvis, Anna Godsmark, George Morris, Bobby Mallick, Phil Eeles. Glasgow Lighthouse Laboratory: Jodie Hay, Harper VanSteenhouse.

Department of Health and Social Care: Jessica Lee.

Welsh Government: Sean White, Tim Evans, Lisa Bloemberg.

Scottish Government: Katie Allison, Anouska Pandya, Sophie Davis.

Public Health Scotland: David I Conway, Margaret MacLeod, Chris Cunningham.

