## Supplementary Material for "Omicron-associated changes in SARS-CoV-2 symptoms in the United Kingdom"

### Supplementary Methods

#### Choice of negative visits in the comparator group

We excluded all negative visits:

1. **From -90 days before** the first S-antibody positive blood test in the study prior to vaccination, where such antibody results are likely to represent previous undetected infection (these results were available only in a random subset of the population);
2. **From -35 days before** the first swab positive onwards from individuals who ever tested PCR positive in the study or positive on either PCR or lateral flow device (LFD) in the linked English testing programme (to avoid ongoing long COVID symptoms[1] and COVID-related symptoms occurring shortly before the positive study test);
3. **From -35 days before** any self-reported positive swab test result onwards (for the same reason; reflecting the fact that individuals may have obtained tests elsewhere)
4. From a small number of individuals who reported either loss of taste or loss of smell at their first study visit and had no national testing programme result within [-21,+21] days (all before 1 July 2020), given the high specificity of this symptom for COVID-19 infection, the fact that it would have been impossible for these individuals to get an external test at the time and the potential for subsequent symptoms to represent long COVID;
5. Where participants reported self-isolating OR contact with **definite** positives in the preceding 28 days (since these individuals have much higher risk of SARS-CoV-2 infection which may not have been detected) and the **previous and the next visit** (because of higher risk of unidentified positivity, and because they may have been contact traced through the national training programme they may be more likely to report symptoms through recall bias, regardless of status);
6. Occurring within [-7,+14 days] of either first, second or third vaccination date[2], to avoid the inclusion of common symptoms caused by vaccination in the test-negative comparator group and to reflect the possibility of small inaccuracies in reported date of vaccination for some participants.

Time windows were arbitrary but aligned with other analyses or windows for considering symptoms associated with PCR-positive episodes.

#### Definition of PCR-positive episodes

#### Historically a new PCR-positive infection “episode” was arbitrarily defined in the study as a new PCR-positive occurring ≥120 days after an index positive with a preceding negative test, or after 4 consecutive negative tests, with 120 days chosen to avoid long intermittent PCR-positivity at high cycle threshold (Ct) values mistakenly being attributed to re-infections[3]. However, re-infections with the Omicron variants occur at much higher frequencies over much shorter time periods compared with earlier variants. Updated definitions of a new PCR-positive episode[4] reflect increasing plausibility of a new infection with longer times from the index positive test, with higher numbers of intervening negative tests, with low Ct values at subsequent positives and with incompatible sequencing data or gene positivity patterns given circulating variants. Following[4], a new PCR-positive infection episode was therefore defined by a new positive test 120 or more days after an initial first positive test and following one or more negative tests, a new positive test 90 or more days after an initial first positive test and following two or more negative tests (or one or more negative tests for new positive tests on or after 20 December 2021, when Omicron BA.1 became the main variant), a new positive test 60 or more days after an initial first positive test and following three or more negative tests, a new positive test after an initial first positive test and following four or more negative tests, a new positive test with evidence, based on genetic sequencing data or PCR gene positivity of a different variant to the index positive, or where both the index/prior positive test and new positive test had a high viral load (low Ct), or there was a large increase in viral load (drop in Ct) between them. We included study tests and tests from the linked English testing programme to define infection episodes (to enable more accurate classification of re-infections), but only used the first study PCR-positive within each infection episode in analyses, since symptoms were elicited only at study visits.

#### Choice of timeframe to include symptoms in the PCR-positive group

Tests are conducted in the survey independently of symptoms, and therefore infection episodes may be identified either early (pre-symptomatic) or late (post-symptomatic). Symptom questions relate to the previous 7 days, so to ensure that subsequently reported symptoms in pre-symptomatic cases were counted we included all symptoms reported at any visit (PCR-positive/PCR-negative/failed) up to 35 days after the index positive test in each infection episode, reflecting the monthly visit schedule.

#### Generalised additive models

In regression models for reporting any evidence of symptoms and specific symptoms in symptomatic PCR-positives and symptomatic PCR-negatives, we truncated age at 85y to avoid undue influence of outliers. Age was modelled as smoothing spline.

bam(cbind(n_withsymptom, n_withoutsymptom) ~

s(study_day, bs="bs", k=75, by=Sars_COV_2_positivity) +

s(age_at_visit, bs="bs", k=15, by=Sars_COV_2_positivity) +

sex:Sars_COV_2_positivity + ethnicity_wo:Sars_COV_2_positivity + sex + ethnicity_wo + Sars_COV_2_positivity, family=binomial(link="cloglog"), method = "fREML", data = data, discrete=TRUE, nthreads =12)

Between 29 September 2021-23 April 2022, including an interaction between study day and age:

bam(symptom_count ~

te(study_day, age_at_visit, bs="bs", k=c(15,15), by=Sars_COV_2_positivity

) +

sex:Sars_COV_2_positivity +ethnicity_wo:Sars_COV_2_positivity + sex + ethnicity_wo + Sars_COV_2_positivity, family=gaussian, method = "fREML", data = data, discrete=TRUE, nthreads =12)

Lines are drawn on all plots at the beginning of the first week during which, based on gene positivity patterns, >50% of PCR-positives were Alpha-compatible (i.e. S-gene negative) 20 December 2020, Delta-compatible (i.e. S-gene positive) 6 June 2021, Omicron BA.1-compatible (i.e. S-gene negative) 19 December 2021, and Omicron BA.2-compatible (i.e. S-gene positive) 27 February 2022.

#### Logistic regression

To assess the association of each symptom with infection in symptomatic PCR-positives vs symptomatic PCR-negatives, we used logistic regression restricted to those reporting any evidence of symptoms. As for the analysis on differences by vaccination status and infection/re-infection, we restricted symptomatic PCR-positives and symptomatic PCR-negatives to those occurring after 29 September 2021 and classified infections as described in the main Methods. For analysis of each variant, PCR-negative visits were restricted to the matching time periods, so the comparator for Omicron BA.1-compatible infections were all symptomatic PCR-negative visits occurring after 1 December 2021 (41,586 visits), for Delta-compatible infections symptomatic PCR-negative visits occurring between 29 September 2021-2 January 2022 (42,740 visits), and for Omicron BA.2-compatible infections all symptomatic PCR-negative visits occurring between 30 January 2022-23 April 2022 (23,076 visits). The outcome was SARS-CoV-2 positivity and the explanatory variables were all 16 symptoms, age, sex and ethnicity (as for the generalised additive models).

model <- bam(SARS-CoV-2_positivity ~

fever + muscle_ache_myalgia + fatigue_weakness + sore_throat + cough + shortness_of_breath + headache + nausea_vomiting + abdominal_pain + diarrhoea + loss_of_taste + loss_of_smell + more_trouble_sleeping + runny_nose_sneezing + wheezing + loss_of_appetite +

sex + ethnicity_wo + s(age_at_visit, bs = "bs", k = 15),

family = binomial, data = dataset_both, method="fREML", discrete = TRUE, nthreads = 12)

Supplementary Tables

**Table S1 Total number of visits per participant over the whole study**

| Number of visits | Number of participants | Percentage of participants |
| --- | --- | --- |
| 1 | 7284 | 1.36 |
| 2 | 3349 | 0.63 |
| 3 | 4043 | 0.75 |
| 4 | 6342 | 1.18 |
| 5 | 13731 | 2.56 |
| 6 | 8704 | 1.62 |
| 7 | 11311 | 2.11 |
| 8 | 13993 | 2.61 |
| 9 | 18625 | 3.48 |
| 10 | 19946 | 3.72 |
| 11 | 19821 | 3.70 |
| 12 | 20841 | 3.89 |
| 13 | 18230 | 3.40 |
| 14 | 20295 | 3.79 |
| 15 | 24349 | 4.54 |
| 16 | 25311 | 4.72 |
| 17 | 28674 | 5.35 |
| 18 | 35525 | 6.63 |
| 19 | 40196 | 7.50 |
| 20 | 41787 | 7.80 |
| 21 | 46211 | 8.63 |
| 22 | 41576 | 7.76 |
| 23 | 30234 | 5.64 |
| 24 | 17201 | 3.21 |
| 25 | 8525 | 1.59 |
| 26 | 5297 | 0.99 |
| 27 | 3929 | 0.73 |
| 28 | 382 | 0.07 |
| >=29 | 25 | 0.00 |

Note: not all these study visits were included in analysis, see Methods and Supplementary Methods.

**Table S2 Number of infections by variant and vaccination status**

| With evidence of symptoms/total (%) | Unvaccinated | First vaccine | Second vaccine | Third vaccine | Fourth vaccine |
| --- | --- | --- | --- | --- | --- |
| Delta | 2696/4753 (57%) | 392/642 (61%) | 5371/7792 (69%) | 389/804 (48%) | 0/0 |
| Omicron BA.1 | 2577/5239 (49%) | 943/1640 (58%) | 3595/6037 (60%) | 13187/21562 (61%) | 3/8 (38%) |
| Omicron BA.2 | 1748/3325 (53%) | 361/637 (57%) | 1549/2630 (59%) | 18839/2784 (67%) | 80/144 (56%) |

### Supplementary Figures

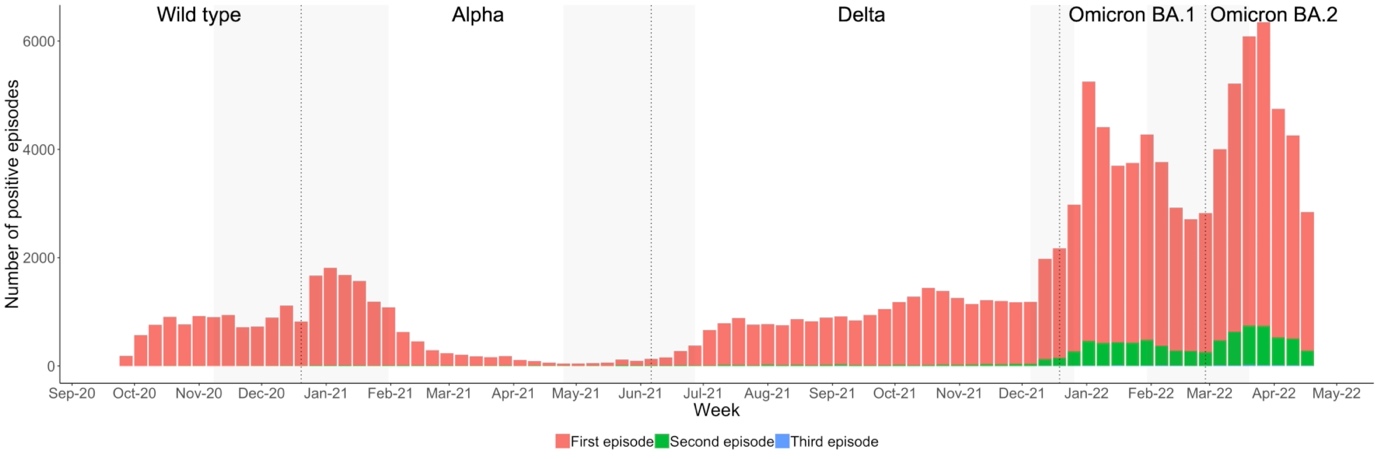

**Figure S1*.* Number of PCR-positive infection episodes by first/second/third episode.**

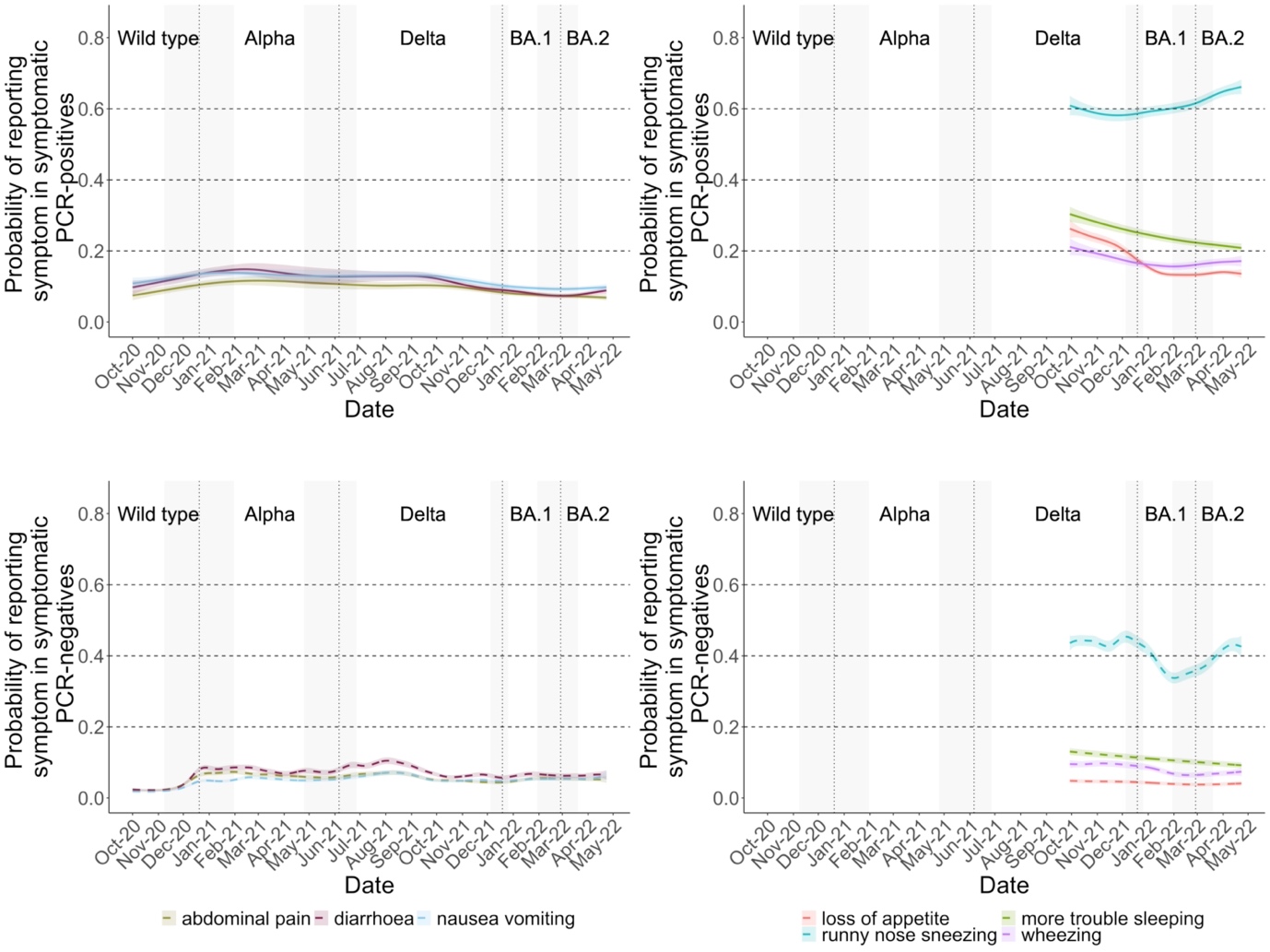

**Figure S2*.* Additional symptoms in those testing positive and negative for SARS-CoV-2 before and following the dominance of Omicron BA.1 in December 2021 and Omicron BA.2 in February 2022 in the UK.** Note: vertical lines indicate periods when new variants came to dominate based on gene positivity patterns, >50% of PCR-positives were Alpha-compatible (i.e. S-gene negative) 20 December 2020, Delta-compatible (i.e. S-gene positive) 6 June 2021, Omicron BA.1-compatible (i.e. S-gene negative) 19 December 2021, and Omicron BA.2-compatible (i.e. S-gene positive) 27 February 2022. Models adjusted for age, sex, ethnicity (presented at the reference category age 45, male, white).

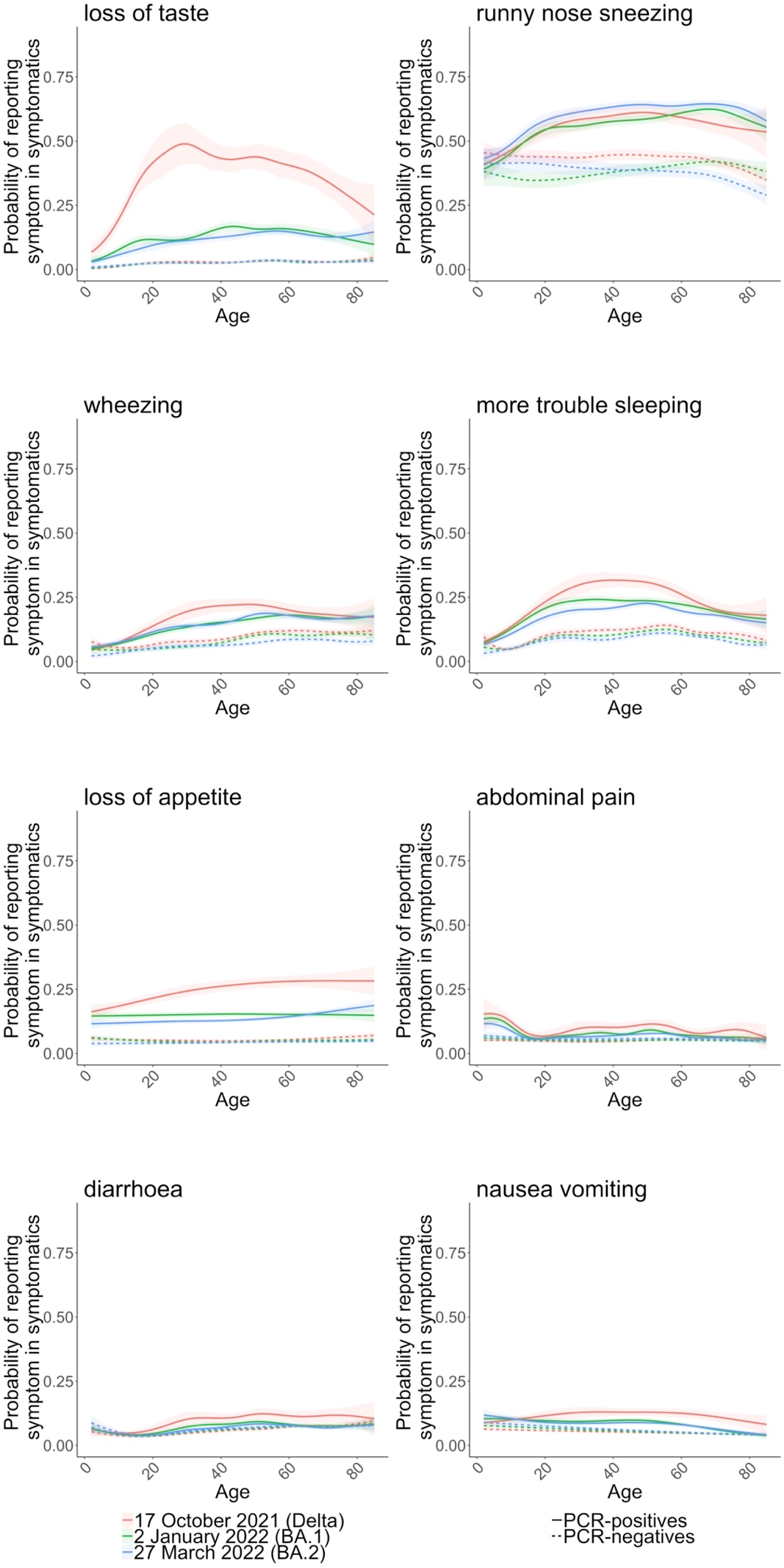

**Figure S3*.* By age, estimated percentage of symptomatic PCR-positives and symptomatic PCR-negatives reporting additional specific symptoms, on 17 October 2021 (beginning of the week with the highest number of Delta-compatible infections), 2 January 2022 (when Omicron BA.1-compatible infections represented the highest proportion of PCR-positives), and 27 March 2022 (when Omicron BA.2 was the dominant variant)**. Panels show the probability of reporting additional specific symptoms in symptomatic PCR-positive infection episodes and in symptomatic PCR-negative comparator study visits from 29 September 2021 onwards at three reference categories, 17 October 2021, 2 January 2022, and 27 March 2022; other symptoms shown in **Fig.4**. All models are adjusted for calendar date, age (allowing for effect modification by calendar date by including an interaction between calendar date and age), sex (reference category male), ethnicity (reference category white).

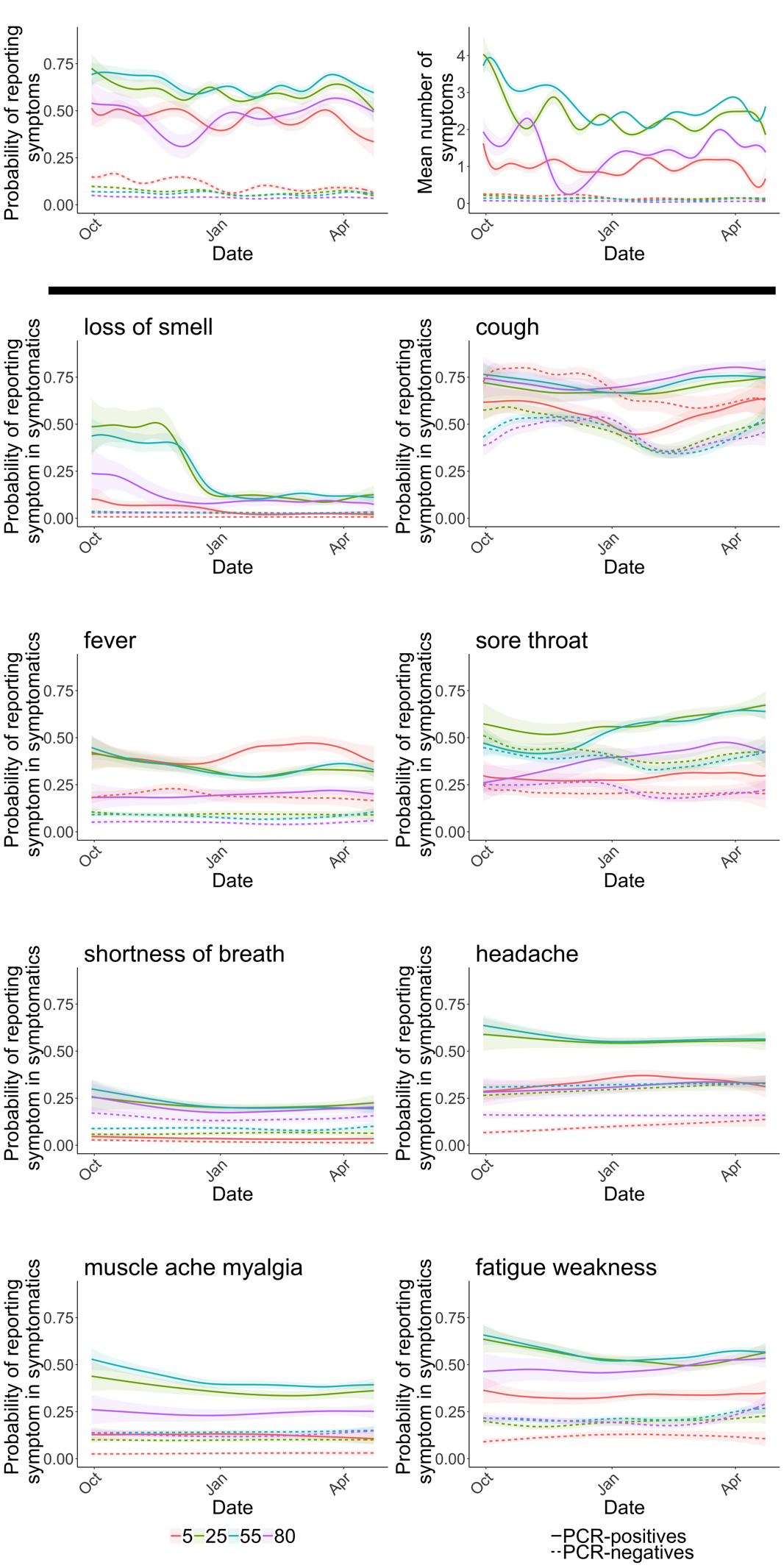

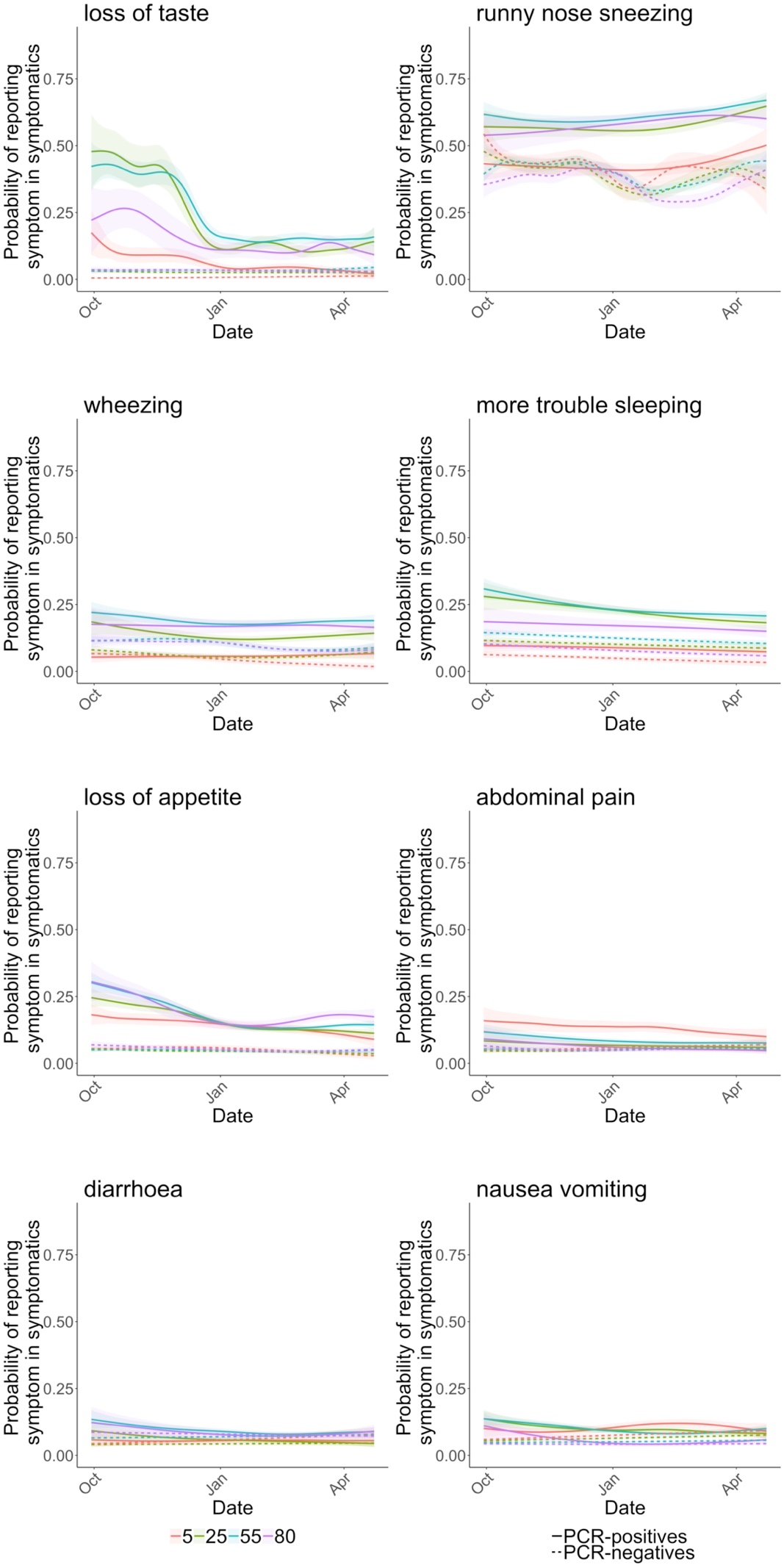

**Figure S4*.* Estimated percentages of those symptomatic and testing positive and negative for SARS-CoV-2 at ages 5, 25, 55, 80, reporting specific symptoms over time**. The panels in the first row show the probability of reporting symptoms and the number of symptoms (out of the 12 elicited throughout the study period) of all PCR-positive infection episodes and all PCR-negative comparator visits at five reference categories, ages 5, 25, 55, 80. The remaining panels show the probability of reporting specific symptoms in symptomatic PCR-positive infection episodes and in symptomatic PCR-negative comparator study visits at these five reference categories, after adjustment for calendar date (allowing for effect modification by age by including an interaction between calendar date and age), age, sex, ethnicity (modelled from 29 September 2021 and presented at reference categories male, white).

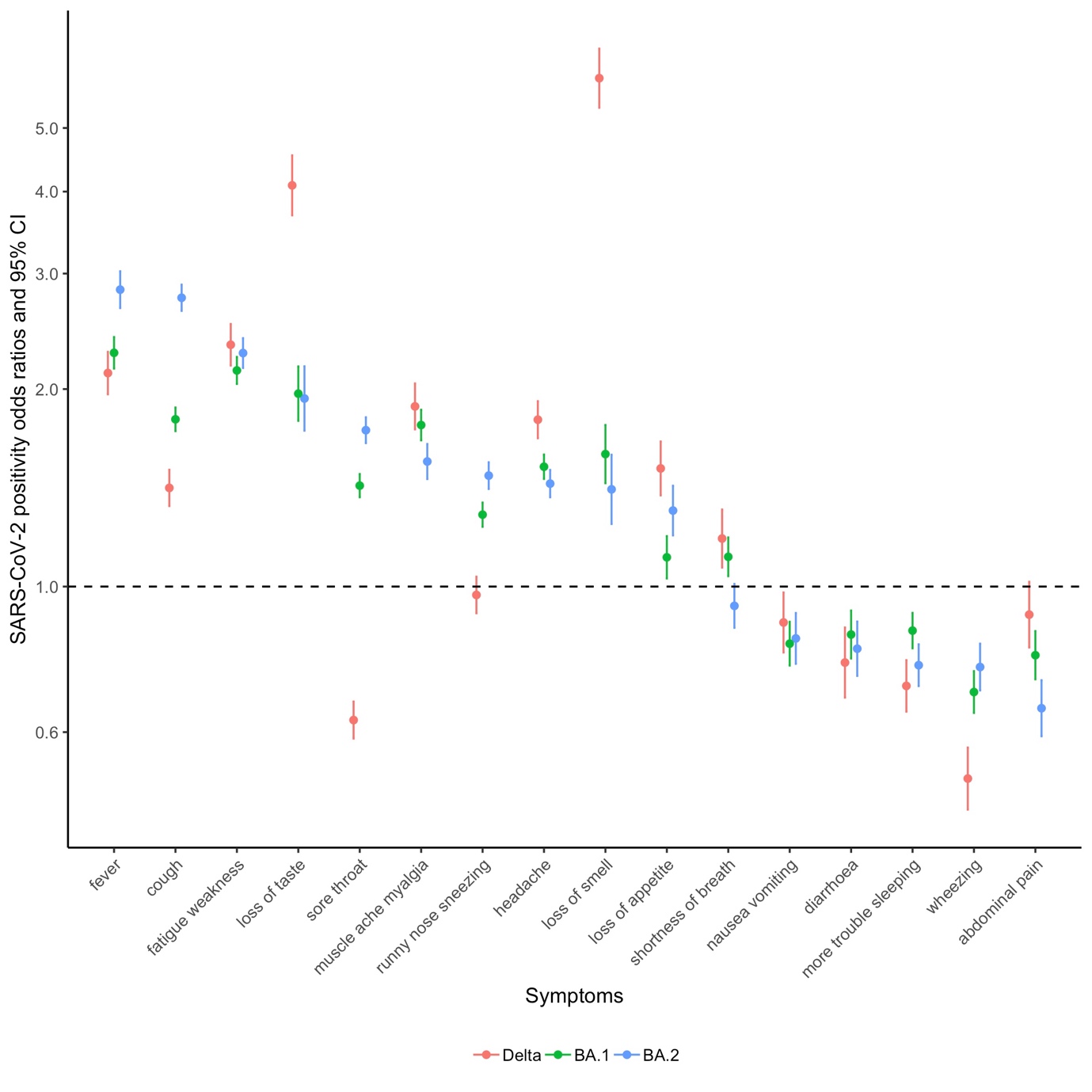

**Figure S5*.* Association between specific symptoms and SARS-CoV-2 positivity with Delta-compatible (S gene positive, 29 September 2021-2 January 2022), Omicron BA.1-compatible (S gene negative, 1 December 2021-23 April 2022) and Omicron BA.2-compatible (S gene positive, 30 January 2022-23 April 2022) infections during matched calendar periods.** From logistic regression models with 16 elicited symptoms as the explanatory variables, mutually adjusted for age, sex and ethnicity.
